# Individualized Atypicality of Structure-Function Configuration Marks Subthreshold Depression in Adolescents

**DOI:** 10.64898/2026.11.20.26368597

**Authors:** Ruisi Wang, Qiwei Guo, Zhongli Lan, Shuai Wu, Weiyin Vivian Liu, Chantat Leong, Wenyue Gao, Jingzhe Zeng, Xinglin Zeng, Andriy Myachykov, Jun Chen, Zhen Yuan

**Author notes:** Corresponding authors: Zhen Yuan,; Jun Chen,; Andriy Myachykov. These authors contributed equally to this work (Ruisi Wang, Qiwei Guo, Zhongli Lan). These authors jointly supervised this work (Andriy Myachykov, Jun Chen, Zhen Yuan).

## Abstract

**Background:** Subthreshold depression (StD) in adolescence represents a critical state preceding major depressive disorder, yet its neurobiological substrates remain poorly characterized.

Conventional structure-function coupling rely on DTI tractography, which produces non-isomorphic matrices and unstable single-subject estimates, limiting their clinical utility. We developed a novel isomorphic and multi-omics framework to quantify individualized structure-function configurations and tested whether adolescents with StD show reproducible deviations from a group-mean reference in a validation sample.

**Methods:** We integrated five structural (cortical thickness, mean curvature, sulcal depth, fractional anisotropy, mean diffusivity) and five functional (ReHo, ALFF, CBF, local and global connectivity strength) features to construct regional SIND-FIND configurations using symmetric KL divergence in a discovery-validation design (N = 131). Individual typicality scores were defined as the correspondence between each participant’s regional configuration profile and HC reference (Leave one out). The score has undergone parallel validations using conventional SC-FC coupling, molecular alignment (PET/AHBA), and behavioral correlations.

**Results:** Group-mean configuration maps were spatially correlated across samples (ρ = .93, p_spin_ < .001). StD participants showed lower typicality than HC (F_(1,94)_ = 8.73, p = .004); the conventional SC–FC comparison was not significant (F_(1,92)_ = 3.31, p = .072). Regional differences involved frontoparietal and salience regions and aligned with PET (r = .55, p_spin_ < .001) and AHBA (r = .47, p_spin_ = .003) maps. After covariate adjustment, typicality correlated with SCL-90 (ρ = −.262, q = .013), SDS (ρ = −.234, q = .022), ISI (ρ = −.296, q = .012) and PSQI (ρ = −.288, q = .012; BH-FDR across five scales); coupling tracked depressive but not sleep symptoms (ISI ρ = −.161, q = .124). Within validation StD participants, SCL-90 remained negatively associated with typicality (ρ = −.42, p = .023).

**Conclusion:** Our isomorphic, multimodal framework captures coordinated structures-function deviation in adolescent StD with greater group-effect sensitivity than conventional SC-FC coupling, together with regional specificity, molecular spatial alignment, and behavioral relevance.

Configuration typicality, defined relative to an HC group-mean reference, may serve as an individual-level marker of organizational deviation during adolescence, pending larger, demographic stratified samples.

## Introduction

Subthreshold depression (StD) involves depressive symptoms that persist below the diagnostic threshold for major depressive disorder (MDD) [1, 2]. Adolescence is a period of increased vulnerability to depressive disorders [3, 4]. Previous neuroimaging studies of StD have often examined individual imaging modalities or brain regions [5]. Multimodal measures can characterize relationships between structural and functional brain organization during adolescence [6–9]. These relationships are commonly quantified as coupling between tractography-derived structural connectivity (SC) and functional connectivity (FC). Coupling varies along the cortical hierarchy, with closer correspondence in unimodal sensory and motor regions and greater divergence in transmodal association cortex [8].

DTI-based SC-FC coupling presents three methodological challenges: 1) it systematically underestimates long-range connections, 2) shows limited agreement with anatomical benchmarks such as cortical similarity measures, and 3) uses structural and functional matrices constructed from different signal properties, complicating their direct comparison. Distributional similarity provides a common way to construct networks from structural and functional features [10–12].

Cortical abnormalities differ between adolescent and adult MDD [13], motivating the study of structure-function organization specifically in adolescents with StD.

The structural features were selected to capture cortical geometry and diffusion properties. Cortical thickness measures the distance between the white and pial surfaces; mean curvature describes local surface bending, and sulcal depth describes position within cortical folds. Fractional anisotropy and mean diffusivity describe the directional dependence and average magnitude of water diffusion, respectively [11]. Their joint distributions retain variation within each region for estimating interregional similarity [10].

The functional features capture regional synchrony (ReHo) [14], low-frequency fluctuation amplitude (ALFF), cerebral blood flow (CBF), and short- and long-range functional connectivity strength (LFCS, GFCS) [15]. The developmental coupling of CBF and ALFF in youth motivates their joint inclusion [16]. Using the morphometric inverse divergence (MIND) approach [10], we constructed two networks: structural inverse divergence (SIND) and functional inverse divergence (FIND) from cortical morphology, diffusion-derived microstructure, and intrinsic functional measures. Regional SIND-FIND configuration was defined as the structure-function correspondence between their interregional similarity profiles. We then examined how these configurations deviate from an HC template in adolescents with StD, testing three hypotheses:

Hypothesis 1 (Organizational Template): Healthy adolescent structure–function configuration shows a spatially stable, reproducible pattern that can serve as a reference for individual typicality.

Hypothesis 2 (Group-Level Deviation and Breadth): Adolescents with StD show reduced typicality relative to healthy controls (HC), and the index additionally captures a broader symptom spectrum than conventional SC–FC coupling.

Hypothesis 3 (Symptom Associations and Molecular Correspondence): (a) Individual typicality is negatively correlated with depressive symptom severity, and this association is reproducible in a quasi-independent sample. (b) The regional configuration contrast map is spatially aligned with Allen Human Brain Atlas (AHBA) gene expression and PET neurotransmitter receptor distributions.

## Methods

### Participants and procedure

The adolescent cohort partially overlapped with our previous glymphatic-related MRI study [17]. This study was approved by the Ethics Committee of Zhuhai Hospital of Guangdong Provincial Hospital of Chinese Medicine and the Ethics Committee of the University of Macau. Before enrolment, participants and legal guardians of those under 18 provided written informed consent. Participants were recruited between April 2024 and June 2025. The discovery cohort initially included 107 participants. The subsequent validation symptom analysis included 29 participants with StD; 16 HC participants from the validation cohort provided its reference configuration profile. Both samples were acquired at the same site under the same MRI protocol and inclusion criteria with no overlapping participants. Initial screening used the Self-Rating Depression Scale (SDS)[18], followed by the behavioral assessment and a structured Diagnostic and Statistical Manual of Mental Disorders, Fifth Edition (DSM-5) based diagnostic interview conducted by experienced clinicians. Participants were 15-22 years old and right-handed. StD was defined as an SDS standard score of at least 53 together with at least two but fewer than five depressive symptoms persisting for at least two weeks; participants did not meet DSM-5 criteria for major depressive disorder. HCs had an SDS standard score below 53 and no personal or family history of psychiatric disorders. The analysis procedures are summarized in Figure 1.

**Figure 1.**
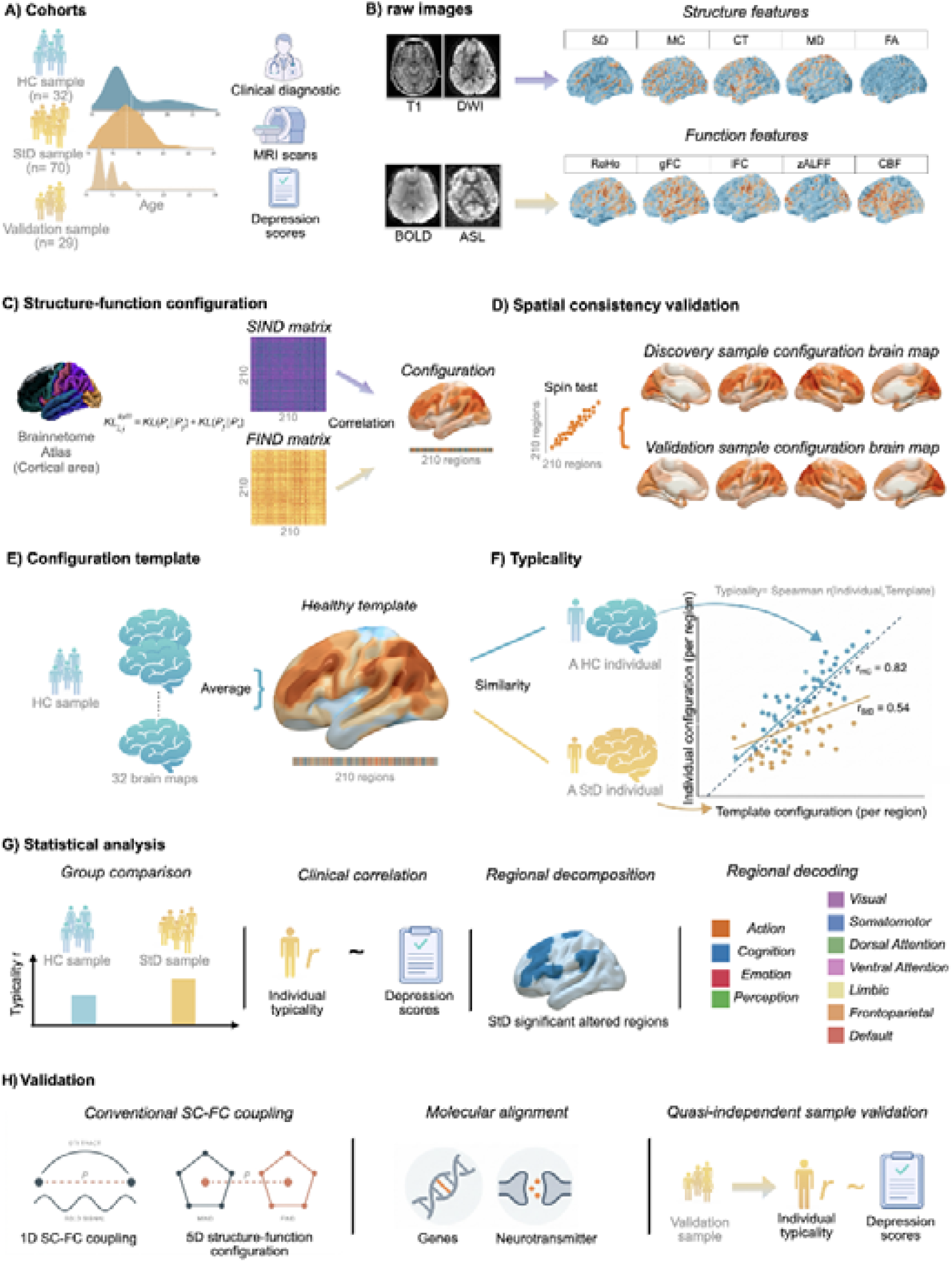
Analysis pipeline. (A) Discovery and validation cohorts with different age distributions; the validation cohort comprised a subsequent sample from the same site. (B) Features derived from multimodal MR sequences (T1, DWI, BOLD, CBF) are grouped into structural (CT, MC, SD, FA, MD) and functional features (ReHo, LFCS, GFCS, ALFF, CBF). (C) Region-wise similarity networks were constructed with a symmetric KL divergence across 210 Brainnetome cortical regions, yielding SIND and FIND matrices; structure-function configuration was defined as the rank correspondence between each region’s structural and functional similarity profiles. (D) Spatial consistency of group-mean configuration maps was assessed across discovery and validation samples with spin testing. (E) An HC configuration template was derived from HC participants in the discovery sample; validation typicality used a separate reference from HC participants in the validation cohort. (F) Individual configuration typicality was quantified as the correlation between each participant’s regional configuration profile and the HC template. (G) Downstream statistical analyses included group comparison, symptom association, regional decomposition, and functional network and behavioral decoding. Additional analyses included conventional SC-FC benchmarking, molecular spatial alignment, and pattern replication in the validation sample. *Note*: HC = healthy controls, StD = Subthreshold depression, CT = cortical thickness, MC = mean curvature, SD = Sulcal depth, FA = fractional anisotropy, MD = mean diffusivity, ReHo = regional homogeneity, ALFF = amplitude of low frequency fluctuation, CBF = cerebral blood flow, LFCS = local functional connectivity strength, GFCS = global functional connectivity strength, KL divergence = Kullback-Leibler, SIND = Structural inverse divergence, FIND = Functional inverse divergence, SC = structural connectivity, FC = functional connectivity.

### Behavioral phenotypes

Depressive symptoms were assessed using the 20-item SDS and the nine-item Patient Health Questionnaire (PHQ-9) [19]. The SDS was used for participant screening, whereas the PHQ-9 indexed dimensional depressive symptom severity. General psychopathological symptoms were assessed with the 90-item Symptom Checklist-90 (SCL-90) [20]. Sleep disturbance was assessed using the seven-item Insomnia Severity Index (ISI) [21] and the 19-item Pittsburgh Sleep Quality Index (PSQI) [22]. Total scores were used to characterize clinical symptoms. The PHQ-9 entered the battery after recruitment had begun and was available for 69 of the 102 discovery participants; Participants with and without PHQ-9 did not differ in age, sex, SDS, SCL-90, ISI or PSQI (all p > .23). The ISI and PSQI were available for 95 participants.

### MRI data preparation

#### Acquisition

MRI data were acquired on a 3.0-T Signa Discovery MR750w scanner (GE Healthcare, Milwaukee, WI, USA) using a 32-channel head coil. T1-weighted images were acquired using a magnetization-prepared rapid gradient-echo sequence (repetition time [TR] = 8.488 ms; echo time [TE] = 3.248 ms; inversion time = 450 ms; flip angle = 12°; field of view [FOV] = 256 × 256 mm²; matrix = 256 × 256; voxel size = 1 × 1 × 1 mm³). Resting-state functional MRI was acquired using gradient-echo echo-planar imaging (TR/TE = 2000/30 ms; flip angle = 90°; FOV = 224 × 224 mm²; matrix = 64 × 64; voxel size = 3.5 × 3.5 × 4.0 mm³; 37 axial slices; 245 volumes). Diffusion-weighted images were acquired using spin-echo echo-planar imaging (TR/TE = 11500/75 ms; flip angle = 90°; FOV = 256 × 256 mm²; matrix = 128 × 128, reconstructed to 256 × 256; voxel size = 2 × 2 × 2 mm³), with 32 non-collinear diffusion directions at b = 1000 s/mm² and one b = 0 image. Perfusion data were acquired using a vendor-specific enhanced ASL sequence with Hadamard-encoded multi-delay labeling using the same 32-channel head coil; TR = 6456 ms, TE = 11.488 ms; flip angle of 111°; 128 × 128 reconstruction matrix; 36 slices; and reconstructed voxel size= 1.7188 × 1.7188 × 4.5 mm³. Seven delay points were acquired with an initial post-labeling delay of 700 ms and a labeling-duration parameter of 3500 ms. The acquisition used four spiral arms and an in-plane parallel-imaging reduction factor = 2.

#### Structural and diffusion image preparation

T1-weighted images underwent cortical surface reconstruction using FreeSurfer 8.1.0 [23], yielding vertex-wise cortical thickness (CT), mean curvature (MC), and sulcal depth (SD). Diffusion-weighted images were processed with MRtrix3 v3.0.8 and FSL v6.0.7.22. Diffusion data underwent denoising (dwidenoise), Gibbs-ringing removal (*mrdegibbs*), and brain extraction from the mean b = 0 image (BET; fractional intensity threshold = 0.25). Motion- and eddy-current-related distortions were corrected using *eddy_cuda* with an anterior–posterior phase-encoding direction and a total readout time of 0.046872 s. Diffusion tensor fitting (*dtifit*) generated fractional anisotropy (FA) and mean diffusivity (MD) maps. FA maps were linearly registered to the JHU-ICBM-FA 1-mm template using FLIRT (12 degrees of freedom; mutual-information cost function), and the resulting transform was applied to MD maps.

#### Functional and perfusion image preparation

Resting-state functional MRI was preprocessed in DPABI [24]: the first 10 volumes were discarded, followed by slice-timing correction, realignment, T1-to-functional coregistration, and tissue segmentation. Data were normalized to MNI space at 3 × 3 × 3 mm³ using DARTEL. Data were temporally filtered at 0.01–0.10 Hz before normalization and were spatially smoothed with a 4-mm full-width at half-maximum Gaussian kernel (FWHM). Mean framewise displacement (FD) was derived from rigid-body realignment parameters and included as a covariate in subsequent group models. Vertex-wise regional homogeneity (ReHo), amplitude of low-frequency fluctuations (ALFF), local functional connectivity strength (LFCS), and global functional connectivity strength (GFCS) were derived from the preprocessed data.

ReHo was calculated from the filtered, unsmoothed BOLD time series as Kendall’s coefficient of concordance among each voxel and its 26 immediate neighbours (3 × 3 × 3 voxel neighbourhood). ALFF was derived in the 0.01–0.10 Hz frequency band and projected to the cortical surface for feature integration.

LFCS and GFCS were calculated from voxel-wise Pearson correlations among grey-matter voxels in MNI space. A grey-matter probability mask thresholded at 0.20 was resampled to the functional-image grid. Voxel time series were standardized before correlation calculation, and self-connections were excluded. For each voxel, LFCS was defined as the mean correlation with grey-matter voxels located less than 20 mm away in Euclidean MNI space, whereas GFCS was defined as the mean correlation with voxels located 20 mm or more away, indexing short-range and long-range functional connectivity strength, respectively.

For CBF quantification, two interleaved eASL image series were reconstructed into seven delay-specific perfusion-weighted images, a combined perfusion-weighted image, and a reference image using Hadamard decoding. The timing schedule was derived from an initial delay of 700 ms, a total labeling-duration parameter of 3500 ms, seven delay points, and a linear-exponential mixing weight of 0.5, with one excitation. The resulting delays were approximately 700, 1061, 1439, 1841, 2277, 2767, and 3358 ms. Voxel-wise arterial transit time (ATT) was estimated using a signal-weighted delay method. Negative delay-specific signals were set to zero. A pCASL model generated a weighted-delay curve on a 1-ms ATT grid spanning the delay range; observed weighted delays were clipped to the curve range and converted to ATT by linear interpolation. Slice-wise 3 × 3 median filtering of ATT preceded CBF quantification from the combined perfusion-weighted image. The model used blood and tissue T1 values of 1.6 s and 1.2 s, labeling efficiency of 0.6375, and a blood–brain partition coefficient of 0.9 mL/g. Native CBF maps were sampled at a fractional cortical depth of 0.5 using mri_vol2surf with header-based registration and trilinear interpolation, then resampled from each individual surface to FreeSurfer *fsaverage* using mri_surf2surf.

#### Image Quality Control

Quality checks at the network-construction stage assessed multimodal feature availability, finite feature values, and correspondence between surface vertices and atlas labels. Vertices outside the cortical parcellation or with all feature values equal to zero were removed before feature standardization. Structural and functional matrices were checked for matching regional labels and order, finite values, symmetry, and unit diagonal elements before regional configuration was calculated.

Multimodal completeness was required for inclusion. Participants entered the configuration analysis only if T1-derived morphometry, diffusion-derived FA/MD, resting-state functional maps, and ASL-derived CBF were all available, so that both 210-region SIND and FIND matrices could be constructed. 5 discovery participants were excluded on this complete-case criterion. 2 HC lacked FA/MD maps, 2 HC lacked usable ASL/CBF, and 1 StD lacked ASL due to incomplete acquisitions. After these procedures, 102 discovery participants (70 StD, 32 HC) and all 29 validation StD participants were retained (n = 131).

#### Harmonization of cortical surface features

To enable multivariate integration, all eligible structural, diffusion, functional, and perfusion feature maps were represented at corresponding cortical vertices in *fsaverage* space before IND network construction, and then assigned to the 210 cortical regions of the Brainnetome (BN210) [25]. CT, MC, and SD were directly normalized to standard space. FA and MD maps in JHU-FA template space were projected to the *fsaverage* surface using nilearn.vol_to_surf (radius = 3 mm; linear interpolation).

ReHo, ALFF, LFCS, and GFCS maps were transformed from volumetric fMRI and were also projected to *fsaverage*. Native CBF maps were sampled with header-based registration to the mid-cortical surface at a fractional cortical depth of 0.5 using trilinear interpolation and resampled from the individual surface to *fsaverage* space.

### SIND and FIND network construction

Following the distributional similarity principle of morphometric inverse divergence (MIND) [10], we constructed structural inverse divergence (SIND) and functional inverse divergence (FIND) networks. The SIND network was generated from the following vertex-wise features: CT, MC, SD, FA, and MD. FIND was generated from ReHo, ALFF, CBF, LFCS, and GFCS.

For each participant, each feature was z-standardized across all retained cortical vertices within its modality-specific feature set. For each pair of BN210 regions, the dissimilarity between multivariate vertex-wise feature distributions was estimated using a k-nearest-neighbour estimator of Kullback-Leibler (KL) divergence with k = 2.

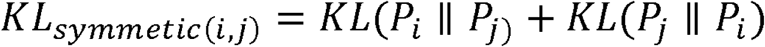

Symmetric divergence was transformed into a bounded similarity value:

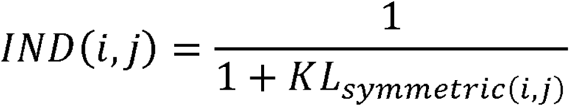

Separate 210 x 210 structural and functional similarity matrices were created for each participant. The diagonal entries were set to 1, and higher off-diagonal values indicated greater similarity between the multivariate feature distributions of two cortical regions. Before structure-function configuration calculation, the structural and functional matrices were required to contain the same 210 BN210 regions in the same order and to have unit diagonal elements. Analyses were restricted to participants with valid structural and functional matrices.

### Structure-function configuration

#### Individual structure-function configuration

Structure–function configuration was defined as the correspondence between structural and functional multivariate similarity profiles. For each of the BN210 regions, the self-similarity element was removed from the corresponding rows of the SIND and FIND network, and Spearman’s rank correlation was calculated between the remaining 209 structural and functional similarity values. This yielded one regional configuration value per cortical region and participant. More positive values indicate greater correspondence between structural and functional similarity profiles.

#### Template of configurations and its validation

The HC reference template was the mean configuration profile across the 210 cortical regions in the discovery HC group. A leave-one-out strategy was adopted: the template for each HC participant was recomputed from the remaining HC participants, whereas each StD participant was compared against the HC template. Cross-sample spatial consistency was assessed by correlating the StD group-mean configuration maps between the discovery and validation samples using Spearman’s rank correlation, followed by a spin test of 5000 rotations of parcellations that preserve spatial autocorrelation [26].

#### Group comparison of individual typicality between StD and HC

For each participant, individual configuration typicality was estimated as the Pearson correlation between that individual’s configuration profile and the HC template, with higher values denoting a more typical configuration. Fisher z-transformed configuration typicality was used for the group comparison by analysis of covariance (ANCOVA), adjusting for sex, age, only-child status, single-parent status, mean framewise displacement (FD) and total intracranial volume (TIV). Effect sizes are reported as partial *η*^2^ and Cohen’s d.

#### Regional and network-level group differences

Regional configuration values were compared between StD and HC at each cortical region using the same covariate set. P values were corrected across regions using Benjamini–Hochberg false discovery rate (BH-FDR). For network-level analyses, regional configuration values were averaged within each of the seven Yeo networks according to a predefined ROI-to-network assignment [27], and the resulting network-level indices were compared between the StD and HC groups with the same covariate-adjusted ANCOVA and BH-FDR correction.

#### Association between configuration typicality and symptom severity

Across the discovery sample, untransformed configuration typicality and each symptom score were separately regressed on age, sex, only-child status, single-parent status, mean FD, and TIV. Two-sided Spearman correlations were then calculated between the residuals, and P values were obtained from the Spearman test applied to these residuals. Each scale used its own complete cases. The identical procedure was applied to typicality derived from conventional SC–FC coupling in the 100 participants with complete coupling estimates, so that the two indices were compared on the same footing. Within each index, BH-FDR correction was applied across the five symptom-scale tests (SCL-90, SDS, PHQ-9, ISI, PSQI).

#### Functional decoding

Brain function decoding was performed based on BrainMap metadata [28]; experiments involving pharmacological interventions or between-group comparisons were excluded to focus on normal physiological conditions. Decoding covered five behavioral domains including cognition, action, perception, emotion, and interoception, each further subdivided into process-specific categories. Regions surviving FDR correction in the regional configuration decomposition were used as the decoding target. Experiments with at least one focus within a spherical search radius of each target region were identified, and a behavioral category was considered associated with that region when the conditional probability of activation given the category exceeded the baseline activation probability [29]. Statistical significance was assessed with a binomial test and corrected across categories using BH-FDR at p < 0.05.

### Validation analyses

#### Conventional structure-function coupling (SC-FC coupling)

Conventional structural-functional connectivity (SC-FC) coupling was computed as a comparison benchmark following a previously described pipeline [30]. SC was defined as the SIFT-2 weighted streamline connectivity between regions, derived from diffusion tractography [31]. FC was defined as the Pearson correlation between regional resting-state BOLD time series. For each region, SC-FC coupling was quantified as the Spearman correlation between that region’s SC profile and FC profile across the remaining regions, yielding a 210-region coupling profile analogous to the SIND-FIND configuration profile. The SC-FC analysis included 100 participants with complete regional and covariate data (69 StD, 31 HC). Its HC reference template and individual typicality scores were constructed after complete-case selection. The group comparison used untransformed typicality r in an ANCOVA with the same six covariates. Network summaries were derived as described above.

#### Gene expression analysis

Gene expression data from six post-mortem donor brains were obtained from the Allen Human Brain Atlas (AHBA) [32] and processed with the abagen toolbox [33]. Transcriptomic profiles were mapped onto the 210 BNA cortical regions. Gene probes were re-annotated, and probes with unreliable gene matches were removed following a standardized imaging-transcriptomics pipeline [34]. Probes detected in fewer than 50% of samples above background were excluded. Tissue samples were assigned to the nearest region within a 2-mm radius based on corrected MNI coordinates. For regions without assigned samples, expression was taken from the sample nearest the regional centroid.

Expression values were sigmoid normalized across donors.

Spatial associations between regional gene expression and the differential configuration t-map were quantified with partial least squares (PLS) regression[35]. The map contained covariate-adjusted contrasts across 210 cortical regions. PLS components maximized covariance between gene expression and this map. The relative variance explained by successive components was quantified, and component significance was assessed with 1000 permutations that randomly reordered regional brain-map values and refitted the PLS model. The leading significant component (PLS1) was retained for interpretation. Spatial correspondence between the fitted PLS1 regional scores and the configuration contrast map was evaluated separately with a spin test. Gene-wise standard errors were estimated by bootstrapping with replacement across regions, and gene Z scores were defined as the ratio of each gene weight to its bootstrap standard error [36].

#### Neurotransmitter receptor analysis

The PET- and SPECT-derived receptor and transporter density profiles were related to the resulting regional configuration contrast map to examine their spatial correspondence. A cortical receptor density map covering serotonin, dopamine, norepinephrine (NET), histamine (H3), acetylcholine, cannabinoid, opioid, glutamate, and GABA systems [37] (github.com/netneurolab/hansen_receptors) was used with an additional FDOPA density map, yielding 21 neurotransmitter markers in total.

Regional estimates were parcellated into the same BNA 210 cortical regions; receptor-transporter profiles and the configuration group effect maps were z-scored across regions.

Spatial associations were quantified with partial least squares (PLS) regression [36]. PLS components were computed as linear combinations of receptor weights that maximally covaried with the configuration group effect map. The cumulative variance explained in the brain map was quantified across components, and component significance was assessed with 1000 permutations that randomly reordered regional values. The leading significant component (PLS1) was retained for interpretation; component weights and scores were sign-flipped for interpretability when the correlation between PLS1 scores and configuration group effect map was negative. Spatial correspondence between the fitted PLS1 regional scores and the configuration contrast map was evaluated separately with a spin test. Receptor-wise standard errors were estimated by bootstrapping with 1000 resamples with replacement across regions. Z scores were defined as the ratio of each weight to its bootstrap standard error. Two-tailed p values were converted from these z scores and corrected using the BH-FDR procedure.

#### Symptom associations in the validation sample

In the validation cohort, configuration typicality was calculated relative to the mean configuration profile of 16 HC participants from that cohort. Symptom associations were evaluated in 29 participants with StD. Typicality and each symptom score were separately regressed on mean FD and TIV within this StD sample, and two-sided Spearman correlations were computed between the resulting residuals. The reported validation p values are unadjusted.

## Results

### Sample characteristics

The discovery sample comprised 102 participants (70 StD, 32 HC), and 29 StD participants formed the validation symptom sample (131 participants across these two analyses); 16 additional HC participants supplied the validation reference profile. Demographic and clinical characteristics are summarized in Table 1 (discovery sample) and in Table 2 (validation sample). The two groups did not differ in age (StD = 17.10 ± 1.35 years, HC = 17.19 ± 2.05 years; Welch’s t_(43.73)_ = −0.22, p = .827) but differed in sex composition, with a higher proportion of females in the StD group (Yates’s 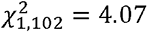, *p* = .044). No region showed a significant main effect of sex after BH-FDR correction. Sex was included as a covariate in all group comparisons.

**Table 1.** Demographic and Clinical Characteristics of the Subthreshold Depression and Healthy Control Groups.

| **Variable** | **StD Group (*n* = 70)** | **HC Group (*n* = 32)** | ***p*** |
|---|---|---|---|
| Age (years) | 17.10 ± 1.35 | 17.19 ± 2.05 | .827 |
| Sex, n (%) |  |  | .044 |
| Male | 23 (32.9) | 18 (56.3) |  |
| Female | 47 (67.1) | 14 (43.8) |  |
| Only child, *n* (%) | 10 (14.3) | 4 (12.5) | .810 |
| Single-parent family, *n* (%) | 9 (12.9) | 2 (6.3) | .268 |
| SDS | 67.96 ± 7.86 | 43.22 ± 6.04 | < .001 |
| SCL-90 | 226.63 ± 72.42 | 118.53 ± 16.17 | < .001 |
| PHQ-9 (n = 37 / 32) | 12.16 ± 5.89 | 1.78 ± 1.36 | < .001 |
| ISI (n = 64 / 31) | 9.80 ± 6.29 | 2.81 ± 3.11 | < .001 |
| PSQI (n = 64 / 31) | 9.09 ± 3.52 | 4.94 ± 2.69 | < .001 |

*Note.* StD = Subthreshold Depression; HC = Healthy Control; SDS = Self-Rating Depression Scale; SCL-90 = Symptom Checklist-90; Continuous variables are presented as *M* ± *SD*. Age, symptom scores, and the binary only-child and single-parent indicators were compared using two-tailed independent-samples *t* tests. Welch's *t* test was reported when Levene's test indicated unequal variances (*p* < .05); Student's *t* test was reported otherwise. Sex distributions were compared using Yates’s continuity-corrected chi-square test. PHQ-9, ISI and PSQI were available for a subset of participants (see n in the respective rows); group comparisons for these scales used Welch's t test.

**Table 2.**
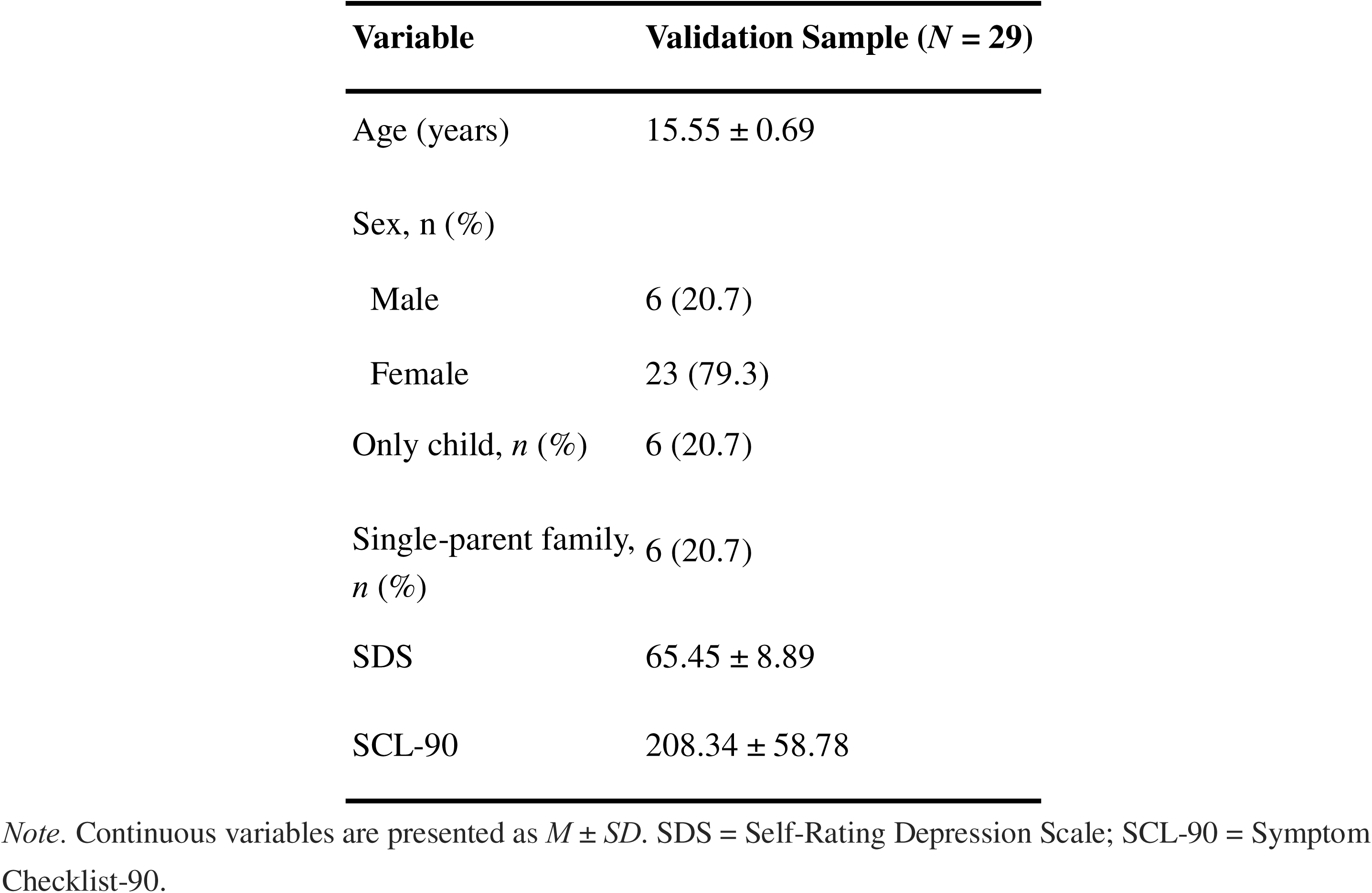
Demographic and Clinical Characteristics of the Validation Sample.

### Spatial consistency of structure-function configuration

The discovery and validation StD samples showed similar group-mean configuration maps (Figure S1A-B), and their group-mean maps were strongly spatially correlated across the cortical regions (*Spearman ρ* = .93, *p_spin_* < 0.001, 5000 rotations; Figure S1C).

### Atypicality of structure-function configuration in StD

Individual configuration typicality was estimated against the healthy configuration template shown in Figure 2A. Adolescents with StD showed significantly lower typicality than HC (*ANCOVA F*_1,94_ = 8.73, *p* = .004, *parital η*^2^ = 0.085, *Cohenn’s d* = − 0.673; Figure 2B), indicating a configuration that departs further from the healthy configuration topography.

**Figure 2.**
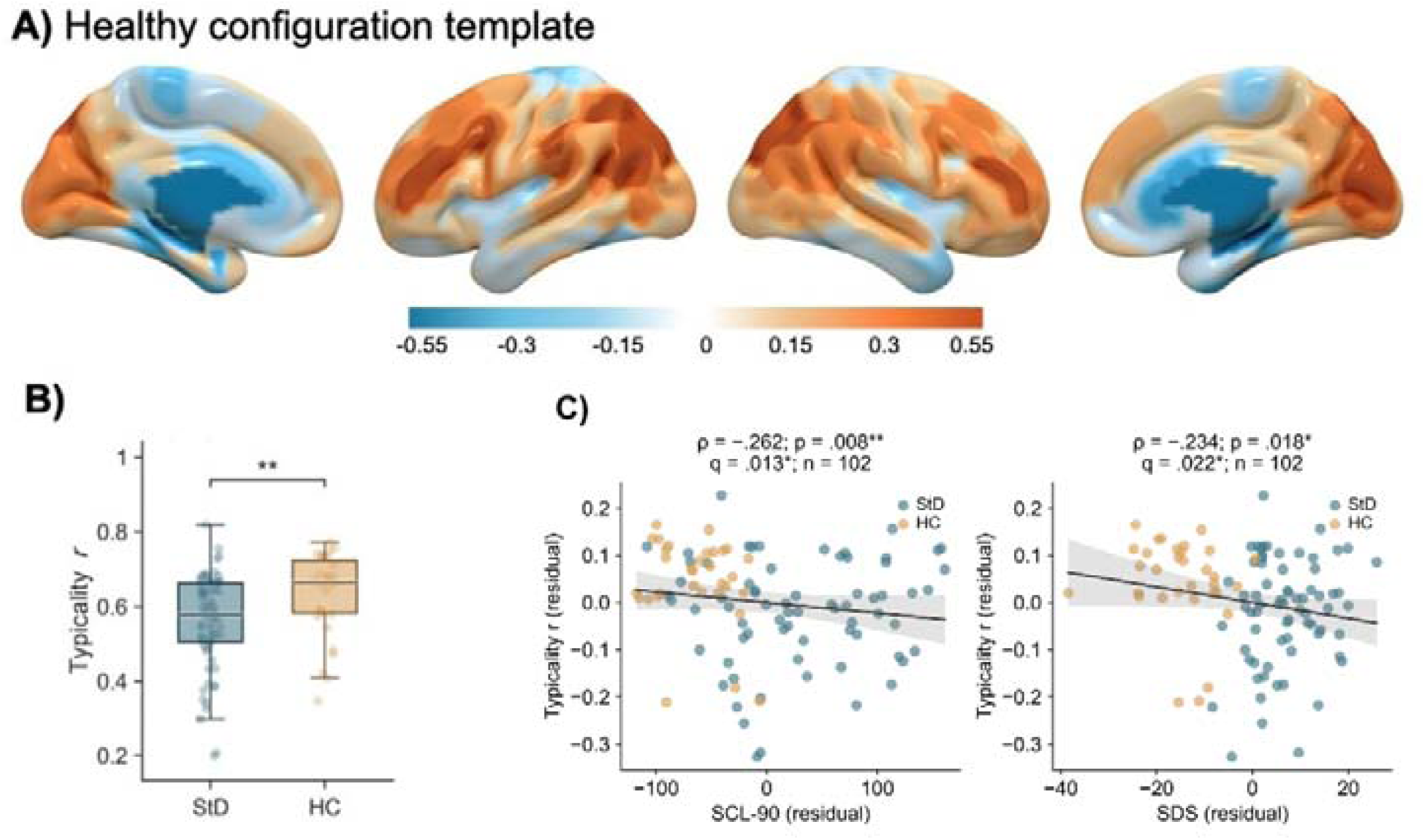
Configuration typicality and symptom associations. (A) HC configuration template on the cortical surface. (B) Untransformed typicality r; the ANCOVA used Fisher z-transformed values and adjusted for age, sex, only-child status, single-parent status, mean FD, and TIV. (C) Typicality and symptom residuals after adjustment for the same six covariates. Annotated Spearman statistics use these residuals. Lines and shaded bands show least-squares fits and their 95% confidence intervals. Note: HC = healthy controls, StD = subthreshold depression, ANCOVA = analysis of covariance, FD = framewise displacement, TIV = total intracranial volume, SCL-90 = 90-item Symptom Checklist, SDS = Self-Rating Depression Scale. In panel B, **p < .01. In panel C, asterisks denote *p < .05 and **p < .01; p denotes the value before multiple-testing correction and q denotes the BH-FDR-adjusted value across the five symptom-scale tests.

### Associations between individual typicality and symptoms

After adjustment for the six covariates, lower configuration typicality was associated with higher scores on every symptom scale (n = 102 for SCL-90 and SDS, 95 for ISI and PSQI, 69 for PHQ-9; Table 3). Within a common BH-FDR correction across the five scales, four survived: SCL-90 (ρ = −.262, p = .008, q = .013), SDS (ρ = −.234, p = .018, q = .022), ISI (ρ = −.296, p = .004, q = .012) and PSQI (ρ = −.288, p = .005, q = .012). The PHQ-9 association was of similar magnitude but did not reach the corrected threshold (ρ = −.236, p = .051, q = .051). The two sleep indices showed the strongest associations of the set (Figure 2C; Table 3).

**Table 3.** Symptom associations of configuration typicality and conventional SC–FC coupling.

| **Symptom scale** | **n** | **Configuration typicality** | **n** | **SC–FC coupling typicality** |
|---|---|---|---|---|
| SCL-90 | 102 | −.262 (.013) | 100 | −.245 (.035) |
| SDS | 102 | −.234 (.022) | 100 | −.228 (.038) |
| ISI | 95 | −.296 (.012) | 93 | −.161 (.124) |
| PSQI | 95 | −.288 (.012) | 93 | −.206 (.060) |
| PHQ-9 | 69 | −.236 (.051) | 68 | −.302 (.035) |

Note. Values are Spearman ρ between covariate-adjusted residuals (age, sex, only-child status, single-parent status, mean FD, TIV), with BH-FDR q across the five scales within each index in parentheses. Sample sizes differ by instrument availability.

### Regional and validation analyses

#### Regional decomposition of altered configuration

Regional ANCOVA identified 9 Brainnetome cortical regions with lower configuration in StD after BH-FDR correction (StD < HC; Figure 3A-B). Of these 9 regions, 4 were in the frontoparietal network: ventrolateral area 8 of right middle frontal gyrus (BNA: A8vl_R; *F* = 17.96, *p_FDR_* = .006), dorsolateral area 8 of right superior frontal gyrus (BNA: A8dl_R; *F* = 16.82, *p_FDR_* = .006), caudal area 40 of right inferior parietal lobule (BNA: A40c_R; *F* = 13.43, *p_FDR_* = .017), and ventral area 4/46 of right middle frontal gyrus (BNA: A9/46v_R; *F* = 10.55, *p_FDR_* = .042); 3 were in the ventral attention/salience network: area 4 of right precentral gyrus (BNA: A4tl_R; *F* = 14.35, *p_FDR_* = .014), area 4 of left precentral gyrus (BNA: A4tl_L; *F* = 12.25, *p_FDR_* = .025), ventral area 44 of right inferior frontal gyrus (BNA: A44v_R; *F* = 10.93, *p_FDR_* = .040); 1 was in the default mode network: caudal area 45 of right inferior frontal gyrus (BNA: A45c_R; *F* = 20.75, *p_FDR_* = .003); 1 was in the somatomotor network: area 4 of right precentral gyrus (BNA: A4t_R; *F* = 10.05, *p_FDR_* = .048) (Figure 3C-D). Detailed regional statistics are provided in the supplementary material. Behavioral decoding of the implicated regions (Figure 3E) linked them to somesthesis/pain, inhibition, execution, attention, and working-memory processes. Specifically, 7 FDR-significant regions were linked to somesthesis/pain in the perception domain.

**Figure 3.**
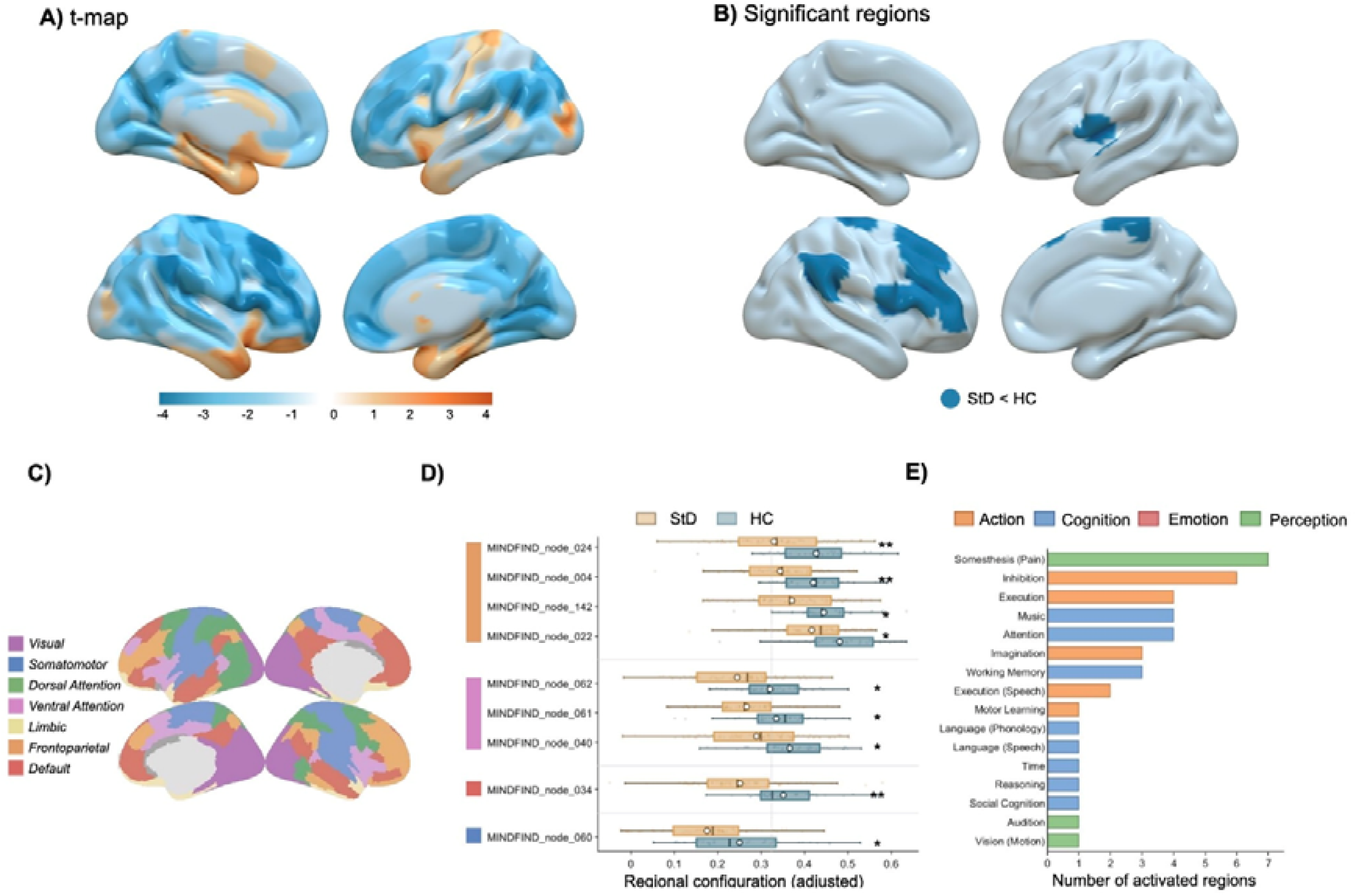
Regional configuration differences and behavioral decoding. (A) Unthresholded StD-HC t map of regional configuration. (B) FDR-significant regions (StD < HC). (C) Yeo seven-network reference. (D) Adjusted regional configuration in FDR-significant nodes (ANCOVA; covariates as in Methods). (E) BrainMap behavioral decoding of FDR-significant regions. Bars show the number of regions associated with each behavioral term, colored by domain (Action, Cognition, Emotion, Perception). **p_FDR < .01; *pFDR < .05.

#### Conventional SC-FC coupling benchmark

In the 100 complete cases (69 StD, 31 HC), typicality derived from conventional SC-FC coupling did not differ significantly between groups (*F*_1,92_ = 3.31, *p* = .072; Figure 4A). Applying the same five-scale procedure to coupling-derived typicality, SCL-90 (ρ = −.245, p = .014, q = .035), SDS (ρ = −.228, p = .023, q = .038) and PHQ-9 (ρ = −.302, p = .012, q = .035) survived correction, whereas the sleep associations did not (ISI ρ = −.161, p = .124, q = .124; PSQI ρ = −.206, p = .048, q = .060). The two indices diverged for sleep rather than for depressive symptoms (Table 3).

**Figure 4.**
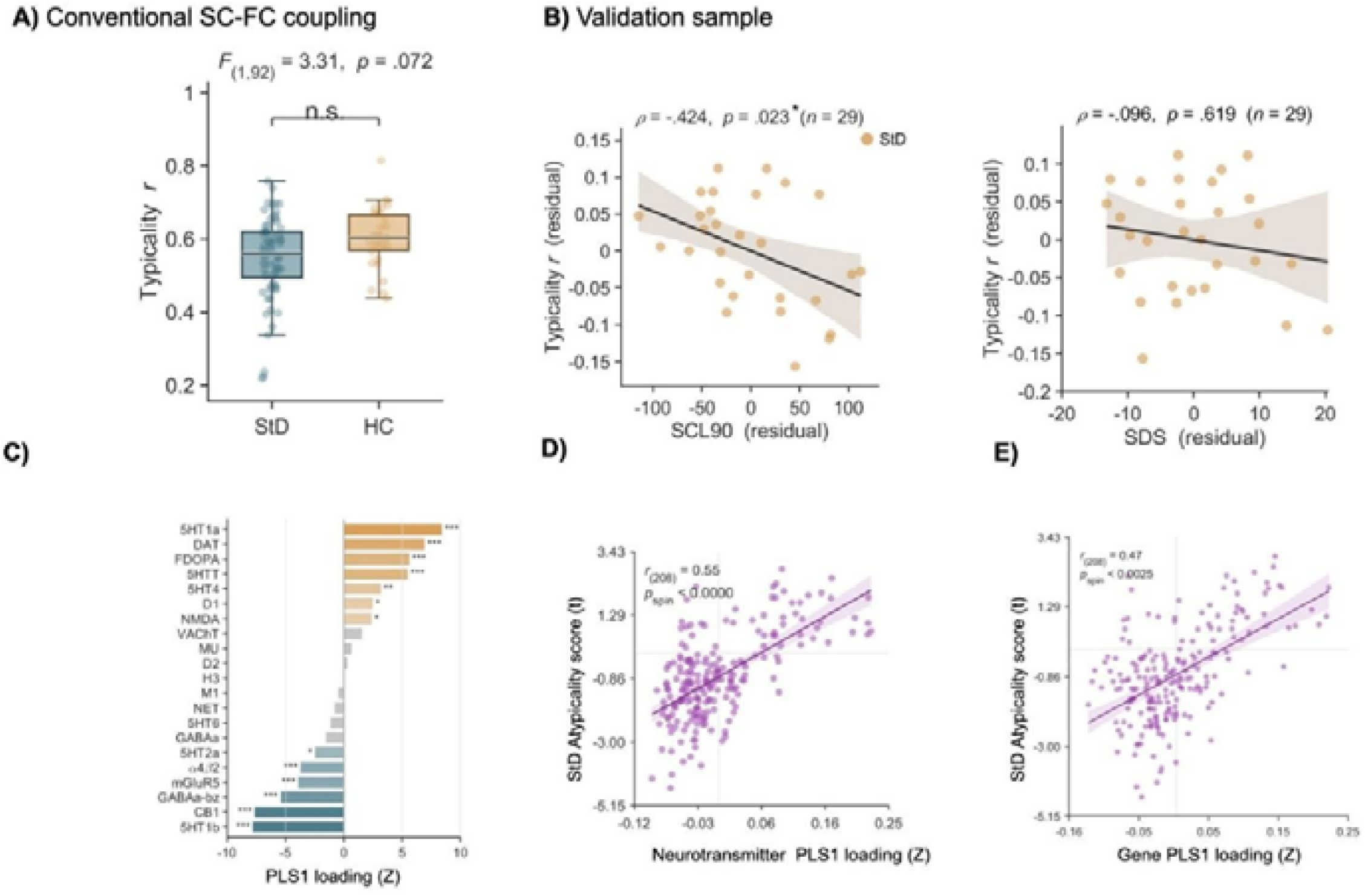
Conventional benchmark, validation-sample symptom association, and molecular spatial alignment. (A) Untransformed typicality r derived from conventional SC-FC coupling in 69 StD and 31 HC participants. (B) Spearman associations between typicality and symptom-score residuals after removing mean FD and TIV in 29 validation StD participants. Typicality was referenced to 16 HC participants in the validation cohort; p values in this panel are unadjusted. (C) Neurotransmitter receptor loadings on the leading PLS component. (D) Spatial correspondence between neurotransmitter PLS1 scores and the regional configuration contrast map. (E) Spatial correspondence between gene-expression PLS1 scores and the same map. In panel C, ***p < .001, **p < .01 and *p < .05 indicate BH-FDR-adjusted receptor-loading tests. Panels D and E report spin-test p values.

#### Symptom associations in the validation sample

In the validation sample (n = 29 StD), lower configuration typicality remained associated with higher SCL-90 score (*ρ* = −.42, *p* = .023), whereas the association with SDS did not reach statistical significance (*ρ* = −.10, *p* = .619) (unadjusted p values; Figure 4B).

#### Molecular spatial correspondence

PLS regression against 21 PET/SPECT neurotransmitter maps yielded a leading component whose regional scores correlated with the regional configuration contrast map (PLS1: *r*_(208)_ = .55, *p_spin_* < 001; Figure 4D), with the strongest positive receptor loadings for 5-HT1a, DAT, FDOPA, 5-HTT, 5-HT4, D1, and NMDA, and the strongest negative loadings for 5-HT1b, CB1, GABAa-bz, and mGluR5 (FDR-corrected; Figure 4C). The parallel PLS analysis of AHBA gene expression showed a comparable spatial alignment (PLS1: *r*_(208)_ = .47, *p_spin_* = .003; Figure 4E).

## Discussion

In this study, we proposed a unified framework that integrates macroscale and microscale features with multimodal functional features to construct structure-function configurations and applied it for the first time in adolescence with StD. Our findings demonstrate that this configuration-based approach captures coordinated structure-functional dysregulation with greater sensitivity than conventional tractography-based SC-FC coupling, and that the observed atypicality is regionally concentrated, molecularly grounded, and behaviorally relevant.

### Stability of the Configuration and Reduced Typicality in StD

Group-mean configuration maps showed consistent cortical topography across the discovery and validation StD samples. The discovery (M = 17.10 ± 1.35 years) and validation (M = 15.55 ± 0.69 years) samples differed in mean age.

Two factors likely underlie this stability. Methodologically, both the SIND (structural) and FIND (functional) matrices were constructed using KL divergence between multivariate regional distributions, achieving isomorphic integration and avoiding the non-isomorphism between conventional SC (DWI tract) and FC (BOLD time series) matrices [10, 38]. Biologically, the stability aligns with the homophily principle, regions with similar structural properties tend to share similar functional profiles, a dominant wiring rule across brain connectomes [11, 12]. Consistent with this, Dorfschmidt et al. reported concordant developmental trajectories between individual structural similarity and functional connectivity networks in adolescence [39]. Together, this regional stability supports the group-level configuration as a reliable "typical template" for individualized typicality estimation.

Based on this template, StD adolescents showed significantly reduced configuration typicality relative to HC, suggesting a deviation from reference structure-function coordination during this sensitive period of cortical remodeling (Hypothesis 2). To benchmark this approach, we constructed a parallel template using conventional DTI tract–based SC–FC coupling. The configuration-based comparison yielded a substantially larger effect (F_1,98_ = 8.73, p = 0.004**) than SC–FC coupling (F_1,92_ = 3.31, p = 0.07), confirming its enhanced sensitivity in detecting coordinated dysregulation in StD.

This enhanced sensitivity likely arises from the multimodal nature of the configuration. Consistent with prior work showing that individualized structural similarity networks outperform conventional SC in characterizing anatomical connectivity [10], the SIND matrix integrates complementary macro- and microstructural features: macrostructural indices (SD, CT, MC) capture cortical morphology, while microstructural indices (MD, FA) reflect underlying tissue organization. This macro–micro integration is critical for constructing biologically meaningful regional similarity, as it jointly encodes cortical geometry and intracortical composition that neither modality alone can fully represent [11, 39]. Analogously, the FIND matrix incorporates multimodal functional features, for instance, CBF and ALFF, which together index neurovascular coupling, a mechanism repeatedly implicated in depression [16]. but poorly captured by conventional FC derived solely from BOLD time-series correlations. Notably, SC-FC coupling also showed a trend-level reduction in StD in the same direction (F_1,92_ = 3.31, p = 0.07) consistent with prior work reporting altered network-based SC–FC coupling in StD relative to HC [40, 41], collectively reinforcing the importance of structure–function relationships in subthreshold depression.

### Regional, Network, and Molecular Basis of Atypicality

Regional decomposition revealed that reduced configuration typicality in StD was not diffusely distributed but converged on nine cortical regions with a clear network-level organization: Four in the FPN, three in the SN, one in the DMN, and one in the SMN. This distribution highlights the FPN and SN as the primary loci of structure–function coordinated dysregulation in adolescents with subthreshold depression.

The predominance of FPN and SN involvement aligns with one of the most consistent findings of meta-analysis in depression which integrated resting-state functional connectivity evidence across 25 studies. The seed-based analysis showed that MDD is characterized by hypoconnectivity within FPN and by disrupted coupling between SN and VAN networks [42]. This convergent pattern across many independent cohorts indicates FPN and SN as core substrate of the disorder. Critically, these alterations are already evident in StD. resting-state studies have reported impaired sub-network of FPN namely cognitive control network and salience-related regions [43, 44]. The present FPN and SN foci fall within this same control hub and SN involved territories, supporting topographical consistency within the function-structure configuration framework.

Behavioral decoding linked these regions to somesthesis/pain, inhibition, execution, attention, and memory. Notably, seven of the nine regions mapped onto somesthesis/pain within the perception domain. This is clinical relevant, as previous evidence indicates that the presence of chronic pain and elevated pain-related symptoms in youth act as a premorbid risk factor for the development of depressive psychopathology in adolescence [45]. The remaining loadings on inhibition, execution, attention, and working memory align with the well-documented cognitive-control deficits of subthreshold and clinical depression [46]. Future studies incorporating direct cognitive and behavioral assessments are warranted to establish the functional consequences of these atypicality.

To test whether the regional configuration atypicality is grounded in the brain’s molecular architecture, we spatially aligned the group-effect configuration map with PET/SPECT neurotransmitter densities and AHBA transcriptomic profiles. Both analyses revealed significant spatial correspondence (PET: r = 0.55, p spin < .001; gene expression: r = 0.47, p spin = .003), indicating that regions showing greater structure–function configuration atypicality in StD co-localize with distinct neurochemical and transcriptomic patterns (Hypothesis 3). This convergence resonates with the homophily principle of brain organization, whereby regions with similar molecular composition tend to develop similar structural and functional profiles [10]. The concordant alignment across two independent molecular modalities suggests that atypical configuration in adolescent subthreshold depression reflects deviation along a molecularly patterned axis of cortical organization rather than stochastic regional variation.

The regional configuration contrast map was spatially associated with PET/SPECT receptor profiles and AHBA gene-expression profiles (PET: r = 0.55, p _spin_ < .001; gene expression: r = 0.47, p _spin_ = .003), as predicted by Hypothesis 3. These associations extend work relating regional molecular composition to cortical similarity [11].

### Behavioral Association and Reproducibility

We observed a negative association between configuration typicality and depressive symptom severity, such that greater deviation exhibited more severe clinical symptoms (Hypothesis 3). As symptom scores in our samples were non-normally distributed (both SDS and SCL-90), spearman correlation was used and the association should be interpreted as monotonic. This negative association suggests that configuration typicality captures a transdiagnostic neural correlate of depressive symptomatology and may serves as an indicator of the graded transition from health to subthreshold and clinical states [47, 48]. The depressive brain-symptom association was further examined in a validation sample. The negative correlation between configuration typicality and SCL-90 was significantly replicated (ρ = -.42, p = .023), while the association with SDS did not reach statistical significance (ρ = -.10, p = .619) but retained the same direction as in the discovery sample. The attenuated SDS effect likely reflects the smaller sample size (n* = 29) and the restricted SDS variability within the purely StD cohort (CV = 13.6% vs. 28.3% for SCL-90), which limits detectable covariance. Given the heterogeneity of depression [48, 49]. prioritizing cross-sample validation is essential for developing neural markers with genuine clinical utility.

Looking ahead, the distributional SIND-FIND framework holds considerable translational promise. Its enhanced stability at the single-subject level relative to conventional SC-FC coupling, positions it as a candidate individual-level signature for early stratification of depression risk during adolescence. Moreover, by enabling direct, symmetric comparison between structural and functional organizational principles, SIND and FIND could help disentangle whether developmental disruptions originate in structural scaffolding, functional dynamics, or their interface-potentially guiding more targeted intervention windows. Future work should test the longitudinal stability of these metrics, their specificity to subthreshold versus clinical depression, and their generalizability across imaging protocols, ages, and diverse populations. Integrating cognitive and behavioral assessments will also be critical to establish the functional relevance of the atypicality detected here.

## Limitations

The StD group had a higher proportion of females. Sex was included as a covariate in all group comparisons, no region showed a significant main effect of sex after correction, and samples with more balanced sex distributions would provide a firmer basis for these associations. Because the SDS served as the screening criterion for group assignment, its association with typicality is partly circular: within-group variance on this instrument is compressed by the screening cutoff, and the SDS estimate is treated as secondary to the SCL-90 result while causal and prognostic inferences, and replication beyond this setting, remain open.

## Conclusion

Configuration typicality captures differences in multimodal brain organization between adolescents with StD and HC. Lower typicality was associated with a broader symptom profile, spanning depressive, general psychopathological and sleep symptoms across the discovery sample, and the association with general symptoms was reproduced within the validation StD sample. Regional configuration differences showed spatial correspondence with cortical molecular maps. This study suggests that structure–function configuration typicality may serve as a sensitive and biologically grounded marker of brain organizational deviation in adolescent StD.

## Data Availability

All data produced in the present study are available upon reasonable request to the authors

## Acknowledgements

We thank the participants and their guardians for taking part in this study, and the radiology staff of Zhuhai Hospital of Guangdong Provincial Hospital of Chinese Medicine for their support with data acquisition. We thank Ms. Ying Mao for helping improve the visual presentation of the manuscript. This work was performed in part at SICC, which is supported by SKL-IOTSC, University of Macau, Macao SAR.

## Data availability

The processed regional feature matrices and individual typicality values supporting the findings are available from the corresponding authors on reasonable request, subject to the ethics approvals under which the data were collected.

---

**Figure S1.**
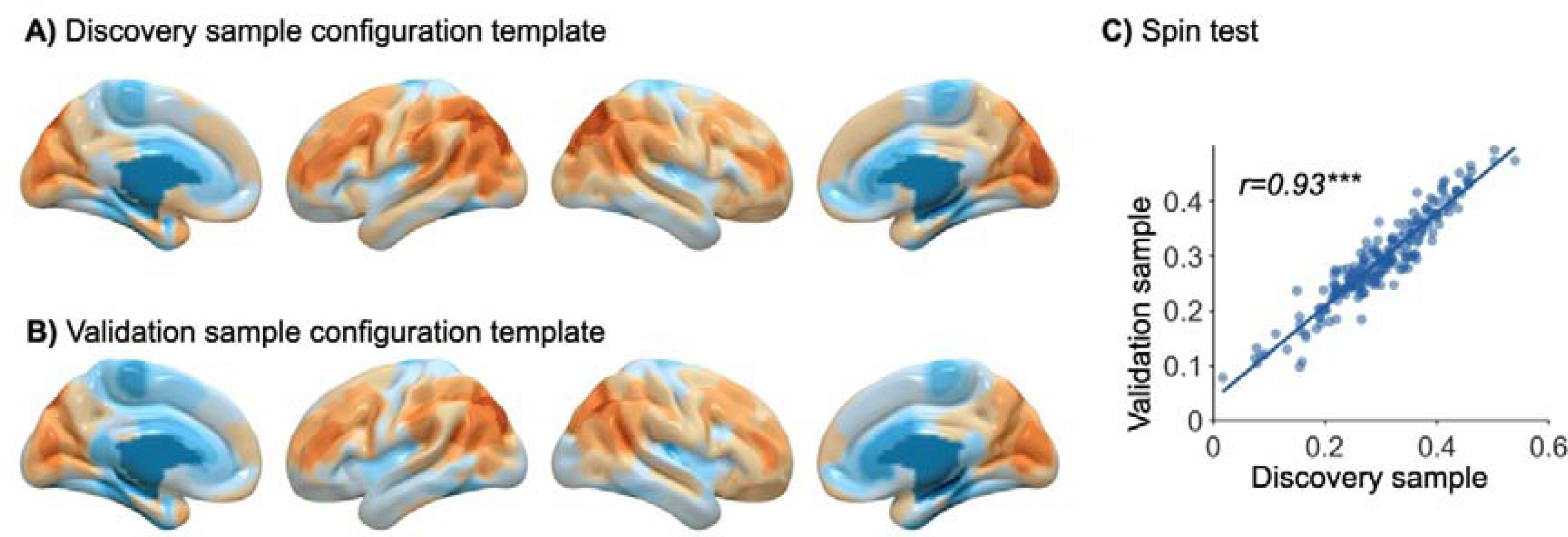
Spatial consistency of StD group-mean configuration across samples. (A) Group-mean cortical distribution of structure-function configuration of StD in the discovery sample and (B) Group-mean cortical distribution of structure-function configuration in the StD validation sample. (C) Spin-test evaluation showing spatial rank correlation between configuration profiles across 210 cortical regions in the discovery sample (n = 70) and the validation sample (n = 29). *Note*: ***p < 0.01, corrected for BH-FDR.

## References

1. Cuijpers P, Smit F. Subthreshold depression as a risk indicator for major depressive disorder: a systematic review of prospective studies. Acta Psychiatrica Scandinavica. 2004;109:325–331.

2. Lee YY, Stockings EA, Harris MG, Doi S a. R, Page IS, Davidson SK, et al. The risk of developing major depression among individuals with subthreshold depression: a systematic review and meta-analysis of longitudinal cohort studies. Psychological Medicine. 2019;49:92–102.

3. Paus T, Keshavan M, Giedd JN. Why do many psychiatric disorders emerge during adolescence? Nat Rev Neurosci. 2008;9:947–957.

4. Salk RH, Hyde JS, Abramson LY. Gender differences in depression in representative national samples: Meta-analyses of diagnoses and symptoms. Psychological Bulletin. 2017;143:783–822.

5. Calhoun VD, Sui J. Multimodal Fusion of Brain Imaging Data: A Key to Finding the Missing Link(s) in Complex Mental Illness. Biological Psychiatry: Cognitive Neuroscience and Neuroimaging. 2016;1:230–244.

6. Gogtay N, Giedd JN, Lusk L, Hayashi KM, Greenstein D, Vaituzis AC, et al. Dynamic mapping of human cortical development during childhood through early adulthood. Proceedings of the National Academy of Sciences. 2004;101:8174–8179.

7. Tamnes CK, Herting MM, Goddings A-L, Meuwese R, Blakemore S-J, Dahl RE, et al. Development of the Cerebral Cortex across Adolescence: A Multisample Study of Inter-Related Longitudinal Changes in Cortical Volume, Surface Area, and Thickness. J Neurosci. 2017;37:3402–3412.

8. Vázquez-Rodríguez B, Suárez LE, Markello RD, Shafiei G, Paquola C, Hagmann P, et al. Gradients of structure–function tethering across neocortex. Proceedings of the National Academy of Sciences. 2019;116:21219–21227.

9. Baum GL, Cui Z, Roalf DR, Ciric R, Betzel RF, Larsen B, et al. Development of structure–function coupling in human brain networks during youth. Proceedings of the National Academy of Sciences. 2020;117:771–778.

10. Sebenius I, Seidlitz J, Warrier V, Bethlehem RAI, Alexander-Bloch A, Mallard TT, et al. Robust estimation of cortical similarity networks from brain MRI. Nat Neurosci. 2023;26:1461–1471.

11. Sebenius I, Dorfschmidt L, Seidlitz J, Alexander-Bloch A, Morgan SE, Bullmore E. Structural MRI of brain similarity networks. Nat Rev Neurosci. 2025;26:42–59.

12. Akarca D, Dunn AW, Hornauer PJ, Ronchi S, Fiscella M, Wang C, et al. Homophilic wiring principles underpin neuronal network topology in vitro. eLife. 2025;14:e85300.

13. Schmaal L, Hibar DP, Sämann PG, Hall GB, Baune BT, Jahanshad N, et al. Cortical abnormalities in adults and adolescents with major depression based on brain scans from 20 cohorts worldwide in the ENIGMA Major Depressive Disorder Working Group. Mol Psychiatry. 2017;22:900–909.

14. Zang Y, Jiang T, Lu Y, He Y, Tian L. Regional homogeneity approach to fMRI data analysis. Neuroimage. 2004;22:394–400.

15. Sepulcre J, Liu H, Talukdar T, Martincorena I, Yeo BTT, Buckner RL. The Organization of Local and Distant Functional Connectivity in the Human Brain. PLoS Computational Biology. 2010;6:e1000808.

16. Baller EB, Valcarcel AM, Adebimpe A, Alexander-Bloch A, Cui Z, Gur RC, et al. Developmental coupling of cerebral blood flow and fMRI fluctuations in youth. Cell Reports. 2022;38:110576.

17. Guo Q, Wang R, Lan Z, Wei Z, Sou U, Zhang C, et al. Integrative Profiling of Glymphatic Dysfunction in Adolescent Subthreshold Depression. 2025:2025.12.18.25342437.

18. Zung WWK. A Self-Rating Depression Scale. Arch Gen Psychiatry. 1965;12:63–70.

19. Kroenke K, Spitzer RL, Williams JBW. The PHQ-9. Journal of General Internal Medicine. 2001;16:606–613.

20. Derogatis LR, Savitz KL. The SCL-90-R, Brief Symptom Inventory, and Matching Clinical Rating Scales. The use of psychological testing for treatment planning and outcomes assessment, 2nd ed, Mahwah, NJ, US: Lawrence Erlbaum Associates Publishers; 1999. p. 679–724.

21. Bastien C. Validation of the Insomnia Severity Index as an outcome measure for insomnia research. Sleep Medicine. 2001;2:297–307.

22. Liu D, Kahathuduwa C, Vazsonyi AT. The Pittsburgh Sleep Quality Index (PSQI): Psychometric and clinical risk score applications among college students. Psychol Assessment. 2021;33:816–826.

23. Fischl B. FreeSurfer. NeuroImage. 2012;62:774–781.

24. Yan C-G, Wang X-D, Zuo X-N, Zang Y-F. DPABI: data processing & analysis for (resting-state) brain imaging. Neuroinformatics. 2016;14:339–351.

25. Fan L, Li H, Zhuo J, Zhang Y, Wang J, Chen L, et al. The human brainnetome atlas: a new brain atlas based on connectional architecture. Cereb Cortex. 2016;26:3508–3526.

26. Alexander-Bloch AF, Shou H, Liu S, Satterthwaite TD, Glahn DC, Shinohara RT, et al. On testing for spatial correspondence between maps of human brain structure and function. NeuroImage. 2018;178:540–551.

27. Thomas Yeo BT, Krienen FM, Sepulcre J, Sabuncu MR, Lashkari D, Hollinshead M, et al. The organization of the human cerebral cortex estimated by intrinsic functional connectivity. Journal of Neurophysiology. 2011;106:1125–1165.

28. Laird AR, Lancaster JL, Fox PT. BrainMap: The Social Evolution of a Human Brain Mapping Database. Neuroinformatics. 2005;3:065–078.

29. Wang R, He Y, Myachykov A, Leong CT, Gao W, Zhang C, et al. Neural-molecular signatures of insomnia: Insights from signed differential mapping and gene expression analysis. NeuroImage. 2026;332:121926.

30. Gu Z, Jamison KW, Sabuncu MR, Kuceyeski A. Heritability and interindividual variability of regional structure-function coupling. Nat Commun. 2021;12:4894.

31. Smith RE, Tournier J-D, Calamante F, Connelly A. SIFT2: Enabling dense quantitative assessment of brain white matter connectivity using streamlines tractography. NeuroImage. 2015;119:338–351.

32. Hawrylycz MJ, Lein ES, Guillozet-Bongaarts AL, Shen EH, Ng L, Miller JA, et al. An anatomically comprehensive atlas of the adult human brain transcriptome. Nature. 2012;489:391–399.

33. Markello RD, Arnatkeviciute A, Poline J-B, Fulcher BD, Fornito A, Misic B. Standardizing workflows in imaging transcriptomics with the abagen toolbox. eLife. 2021;10:e72129.

34. Arnatkevic iūtė A, Fulcher BD, Fornito A. A practical guide to linking brain-wide gene expression and neuroimaging data. Neuroimage. 2019;189:353–367.

35. Whitaker KJ, Vértes PE, Romero-Garcia R, Váša F, Moutoussis M, Prabhu G, et al. Adolescence is associated with genomically patterned consolidation of the hubs of the human brain connectome. Proc Natl Acad Sci U S A. 2016;113:9105–9110.

36. Morgan SE, Seidlitz J, Whitaker KJ, Romero-Garcia R, Clifton NE, Scarpazza C, et al. Cortical patterning of abnormal morphometric similarity in psychosis is associated with brain expression of schizophrenia-related genes. Proc Natl Acad Sci. 2019;116:9604–9609.

37. Hansen JY, Shafiei G, Markello RD, Smart K, Cox SML, Nørgaard M, et al. Mapping neurotransmitter systems to the structural and functional organization of the human neocortex. Nat Neurosci. 2022;25:1569–1581.

38. Suárez LE, Markello RD, Betzel RF, Misic B. Linking Structure and Function in Macroscale Brain Networks. Trends in Cognitive Sciences. 2020;24:302–315.

39. Dorfschmidt L, Váša F, White SR, Romero-García R, Kitzbichler MG, Alexander-Bloch A, et al. Human adolescent brain similarity development is different for paralimbic versus neocortical zones. Proc Natl Acad Sci U S A;121:e2314074121.

40. Suo X, Chen L, Kemp GJ, Wu D, Wang S. Aberrant Structural–Functional Coupling of Large-Scale Brain Networks in Older Women With Subthreshold Depression. J Gerontol B Psychol Sci Soc Sci. 2025;80:gbaf013.

41. He Y, Zhao B, Liu Z, Hu Y, Song J, Wu J. Individualized identification value of stress-related network structural-functional properties and HPA axis reactivity for subthreshold depression. Transl Psychiatry. 2024;14:501.

42. Kaiser RH, Andrews-Hanna JR, Wager TD, Pizzagalli DA. Large-scale network dysfunction in major depressive disorder: A meta-analysis of resting-state functional connectivity. JAMA Psychiatry. 2015;72:603–611.

43. Hwang JW, Egorova N, Yang XQ, Zhang WY, Chen J, Yang XY, et al. Subthreshold depression is associated with impaired resting-state functional connectivity of the cognitive control network. Transl Psychiatry. 2015;5:e683–e683.

44. Peng X, Lau WKW, Wang C, Ning L, Zhang R. Impaired left amygdala resting state functional connectivity in subthreshold depression individuals. Sci Rep. 2020;10:17207.

45. Soltani S, Noel M, Bernier E, Kopala-Sibley DC. Pain and insomnia as risk factors for first lifetime onsets of anxiety, depression, and suicidality in adolescence. Pain. 2023;164:1810–1819.

46. Dotson VM, McClintock SM, Verhaeghen P, Kim JU, Draheim AA, Syzmkowicz SM, et al. Depression and Cognitive Control across the Lifespan: a Systematic Review and Meta-Analysis. Neuropsychol Rev. 2020;30:461–476.

47. Noyes BK, Munoz DP, Khalid-Khan S, Brietzke E, Booij L. Is subthreshold depression in adolescence clinically relevant? Journal of Affective Disorders. 2022;309:123–130.

48. Li J, Long Z, Ji G-J, Han S, Chen Y, Yao G, et al. Major depressive disorder on a neuromorphic continuum. Nat Commun. 2025;16:2405.

49. Winter NR, Leenings R, Ernsting J, Sarink K, Fisch L, Emden D, et al. Quantifying Deviations of Brain Structure and Function in Major Depressive Disorder Across Neuroimaging Modalities. JAMA Psychiatry. 2022;79:879–888.

